# Surgical Revision of Deep Brain Stimulation After Suboptimal Clinical Outcomes: A Systematic Review and Exploratory Meta-analysis

**DOI:** 10.64898/2026.10.31.26366990

**Authors:** Mostafa Abdollahi Sarvi, Saghar Babazadeh

## Abstract

**Background:** Some patients obtain less benefit than expected from deep brain stimulation (DBS) despite optimized programming. We systematically reviewed the outcomes of surgical revision in this setting.

**Methods:** Following PRISMA 2020, we searched PubMed, Embase and Scopus to October 2026 for studies of lead repositioning, replacement, added (rescue) leads or target switching performed for inadequate benefit or stimulation-related adverse effects; revision for infection or hardware failure was excluded. Outcomes were pooled with random-effects models, and certainty was rated with GRADE.

**Results:** Seventeen case series (approximately 149 patients) were included. Across 14 studies (95 patients), 81% (95% CI 68–91%) improved meaningfully, with similar estimates for same-target revision (82%) and for added or switched leads (79%). In Parkinson disease, the off-medication/on-stimulation motor score fell by 15.6 points after lead revision (95% CI −22.6 to −8.6; four studies). Benefit was greatest when a lead outside the dorsolateral subthalamic nucleus was corrected and levodopa response was preserved. Seven studies reported surgical or hardware complications, and two deaths occurred. Certainty of evidence was very low.

**Conclusions:** Most published revised patients improve, but the evidence derives from small uncontrolled series. Revision should follow imaging, hardware checks and exhaustive reprogramming, within multidisciplinary review.

## Introduction

Deep brain stimulation (DBS) is an established treatment for advanced Parkinson disease, essential tremor and dystonia. Its efficacy is supported by randomized trials [1–4], and long-term follow-up shows that motor benefit in Parkinson disease is sustained for many years [5], while indications and technology continue to expand [6]. In routine practice, however, results vary between patients and centers. Experienced centers increasingly receive patients whose outcome fell short of expectation [7,8], and an analysis of more than 28,000 DBS procedures across multiple databases reported revision and removal rates above 15% [7].

A suboptimal outcome can be defined as benefit that falls clearly short of what the preoperative assessment predicted (in Parkinson disease, usually the levodopa response) or benefit that is limited by stimulation-induced adverse effects. Its causes fall into four broad groups: inappropriate patient selection or an incorrect diagnosis, inadequate programming, suboptimal lead position, and loss of benefit over time. In a series of 41 patients referred after apparent DBS failure, 46% had a misplaced lead, 37% were inadequately programmed and 12% had been given the wrong diagnosis; after programming, medication changes and, in some cases, lead replacement, 51% finally did well [8]. In a later German series, 9 of 48 referred patients (19%) remained suboptimal after extensive reprogramming [9]. Hardware faults such as lead fracture, short circuits or disconnection can mimic loss of efficacy and must be excluded before a clinical cause is assumed [10,11].

Lead position matters because DBS targets are small and functionally segregated. The sensorimotor territory of the subthalamic nucleus (STN) occupies its dorsolateral portion and lies close to the internal capsule, the medial lemniscus and fibers of the oculomotor system. A lead displaced by a few millimeters may therefore stimulate associative or limbic territories, or recruit corticobulbar and corticospinal fibers at low amplitudes, so that the therapeutic window closes before adequate benefit is reached. Accordingly, stimulation within the dorsolateral STN on magnetic resonance imaging predicts motor outcome in Parkinson disease [12], and lead location is similarly related to tremor control after thalamic stimulation for essential tremor [13]. Errors of this size can arise from targeting inaccuracy, brain shift after loss of cerebrospinal fluid during surgery, or lead migration.

Benefit may also fade despite a well-placed lead. In essential tremor, progressive loss of efficacy after thalamic stimulation has been attributed to habituation or tolerance, to disease progression, or to both [14,15]. In Parkinson disease, the emergence of axial and other levodopa-resistant symptoms limits what any target can achieve over time [5]. Distinguishing these mechanisms matters clinically: a misplaced lead can be corrected surgically, whereas tolerance or progression may not respond to a new lead placed in the same circuit.

When programming has been exhausted, including newer strategies such as directional current steering and short pulse widths that widen the therapeutic window [16,17], surgeons can reposition or replace the lead, add a second lead in the same or another target (a rescue lead) [18,19], or switch to a different target. Each option involves trade-offs. Repositioning corrects misplacement but risks losing a partial benefit already obtained; a rescue lead preserves the original lead but adds hardware and usually requires a dual-channel generator; and switching target abandons the original circuit altogether. Every repeat procedure also carries the hemorrhage, infection and hardware risks of DBS surgery [20].

Evidence for these options comes from small series that differ in disease, technique and outcome definition, and it has not been synthesized. Most published work on DBS revision concerns infection and hardware failure, which answers a different clinical question [7,20]. To our knowledge, no review has addressed what happens to patients who undergo revision specifically because their clinical result was inadequate. We therefore systematically reviewed the clinical outcomes, complications and predictors of success of surgical revision in patients with suboptimal DBS outcomes despite optimized programming, and compared same-target revision with added, rescue or switched leads.

## Methods

### Protocol, registration and reporting

The review was conducted and reported according to the PRISMA 2020 statement [21]. The protocol was registered in the International Prospective Register of Systematic Reviews (PROSPERO; CRD420261557397, registered 30 October 2026). The review started on 30 August 2026. Differences between the protocol and the review are listed at the end of the Methods.

### Eligibility criteria

We included original clinical studies (randomized trials, prospective or retrospective cohorts, case-control studies and case series of two or more patients) of humans of any age and any DBS indication who had suboptimal outcomes despite appropriate programming or clinical optimization and then underwent surgical revision. Revision comprised lead repositioning, replacement or reimplantation, and we also accepted rescue leads and target switching when outcomes after surgery were reported. Revision had to be done to improve inadequate therapeutic benefit or to resolve stimulation-related adverse effects. Studies had to report at least one of clinical improvement, predictors of success, complications, adverse effects or quality of life, be written in English, and provide enough full-text data for extraction.

We excluded studies of primary implantation without revision, programming changes alone, revision performed only for infection, hardware malfunction, battery depletion or device replacement, ablative procedures, target switching without postoperative outcomes, and purely technical or radiological reports. We also excluded reviews, editorials, letters, commentaries, conference abstracts, animal or laboratory studies, single-patient case reports, duplicate cohort reports and studies without obtainable full text.

### Search and selection

We searched PubMed (MEDLINE), Embase and Scopus from database inception to 1 October 2026, with no date restrictions, and checked the reference lists of included studies (backward citation searching). Only published studies were sought. The PubMed strategy combined controlled vocabulary and free-text terms for deep brain stimulation (Deep Brain Stimulation [MeSH]; “deep brain stimulation”, “deep-brain stimulation” and DBS in titles and abstracts) with terms for revision (Reoperation and Electrodes, Implanted [MeSH]; reoperat*, revis*, reimplant*, reposition*, lead or electrode replacement, lead or electrode revision, surgical revision, lead relocation, target switching, target substitution, rescue surgery and salvage surgery). The strategy was adapted to the syntax of Embase and Scopus; all strategies are reproduced in Supplementary Appendix 1 and were uploaded with the PROSPERO record. Records from the three databases were combined and duplicates removed using Endnote. Two reviewers independently screened titles and abstracts and then full texts against the eligibility criteria, and disagreements were resolved by discussion until consensus. The full-text stage assessed 21 reports.

### Data extraction

We extracted study design, center, country, DBS indication, number of patients and leads revised, revision strategy, indication for revision, interval from the index operation, lead displacement, surgical approach, follow-up, outcome measures and results, complications, and reported predictors of success. Two reviewers extracted data independently using a standardized form, and discrepancies were resolved by discussion. When a study mixed revision for suboptimal outcome with infection or device failure, we extracted only the suboptimal-outcome subgroup. When patient overlap between reports was possible, we kept one report per patient. Study authors were not contacted for missing or unclear information.

### Outcomes

The main outcomes, specified in the protocol, were clinical improvement after revision and complications. Clinical improvement was measured with disease-specific scales (for example UPDRS-III or MDS-UPDRS-III in Parkinson disease, the Fahn–Tolosa–Marin Tremor Rating Scale for tremor and the Burke–Fahn–Marsden Dystonia Rating Scale motor score for dystonia) or with clinician-rated global impression, at the last reported follow-up after revision. Complications comprised surgical and hardware events (such as infection, lead or extension fracture, wound-healing problems and hemorrhage) and stimulation-related adverse effects. Factors associated with success or failure, and differences between same-target revision and added, rescue or switched leads, were examined as secondary questions. For the pooled proportion we defined “meaningful improvement” from each study’s own criteria (for example, marked improvement, a clinical global impression of much or very much improved, or a reduction of at least 25% in motor score), and separately counted “any improvement”. The definitions used for each study are given in Supplementary Table S2a.

### Risk of bias and certainty of evidence

Because every included study was a case series, we used the Joanna Briggs Institute (JBI) checklist for case series [22] and considered the domains proposed by Murad and colleagues [23]. We added judgements on comparator, blinding of outcome assessment, missing data and selective reporting, which the checklist does not capture. Two reviewers appraised each study independently and resolved disagreements by discussion; study investigators were not contacted when information was unclear. Selective reporting and missing data were judged within this appraisal. Certainty of evidence for each outcome was rated with GRADE [24], considering risk of bias, inconsistency, indirectness, imprecision and publication bias. Because formal tests were not feasible, publication bias was judged qualitatively and incorporated into the GRADE rating.

### Statistical analysis

We pooled single-arm data with DerSimonian–Laird random-effects models [25]. Proportions were transformed with the Freeman–Tukey double arcsine method [26,27] and back-transformed for reporting. We analyzed studies of same-target revision (repositioning or replacement) and of added, rescue or switched leads as subgroups. For continuous outcomes we pooled the change in off-medication/on-stimulation motor score (points). Standard deviations of change were calculated from raw patient data when available, derived from reported p values, or imputed assuming a pre–post correlation of 0.5 (tested at 0.3 and 0.7). Medians and ranges were converted to means and standard deviations by the method of Wan and colleagues [28]. Heterogeneity was expressed as I² and τ² [29], and a Hartung–Knapp interval was added for the continuous outcome [30]. Sensitivity analyses comprised leave-one-out analysis, exclusion of studies with fewer than five patients and alternative counting of one study with internally inconsistent data. Publication bias was not tested statistically because of the few, small studies. Studies without patient-level or comparable outcome data were synthesized narratively, as were comparator data reported within studies (for example, reimplantation after infection). Analyses were performed in python 3.14.

### Differences between protocol and review

The review followed the registered protocol. The following were not specified in the protocol: (1) “any improvement” was analyzed alongside meaningful improvement; (2) medians and ranges were converted with the method of Wan and colleagues [28], and a Hartung–Knapp interval was added for the continuous outcome [30]; (3) sensitivity analyses were added (leave-one-out, exclusion of studies with fewer than five patients, alternative counting of one internally inconsistent study and alternative pre–post correlations); and (4) the JBI checklist was supplemented with additional bias domains [23]. As anticipated in the protocol, complications, the essential tremor and dystonia cohorts and predictors of success were synthesized narratively.

## Results

### Study selection and characteristics

The searches of PubMed, Embase and Scopus returned 3,093 records, of which 369 were duplicates. Of the 2,724 unique records screened by title and abstract, 2,339 (85.9%) were indexed as human studies, 226 (8.3%) as animal-only studies and 131 (4.8%) were not yet MeSH-indexed. Case reports (383; 14.1%) and narrative reviews (316; 11.6%) were the most common publication types, followed by other clinical trials (116; 4.3%), randomized trials (51; 1.9%), systematic reviews (46; 1.7%) and meta-analyses (22; 0.8%); 161 records (5.9%) had no abstract and 84 (3.1%) were not in English (Supplementary Table S7). Screening excluded 2,703 records, and all 21 remaining reports were retrieved and assessed in full text. Four were excluded: two examined hardware failure only (electrode fracture or disruption, and short circuits or disconnections) [10,11], one was a general adverse-event series describing few revisions [31], and one was a tractography study in which a single patient was revised [32]. Also, backward citation searching identified no additional eligible studies. Seventeen studies were included (Figure 1).

**Figure 1.**
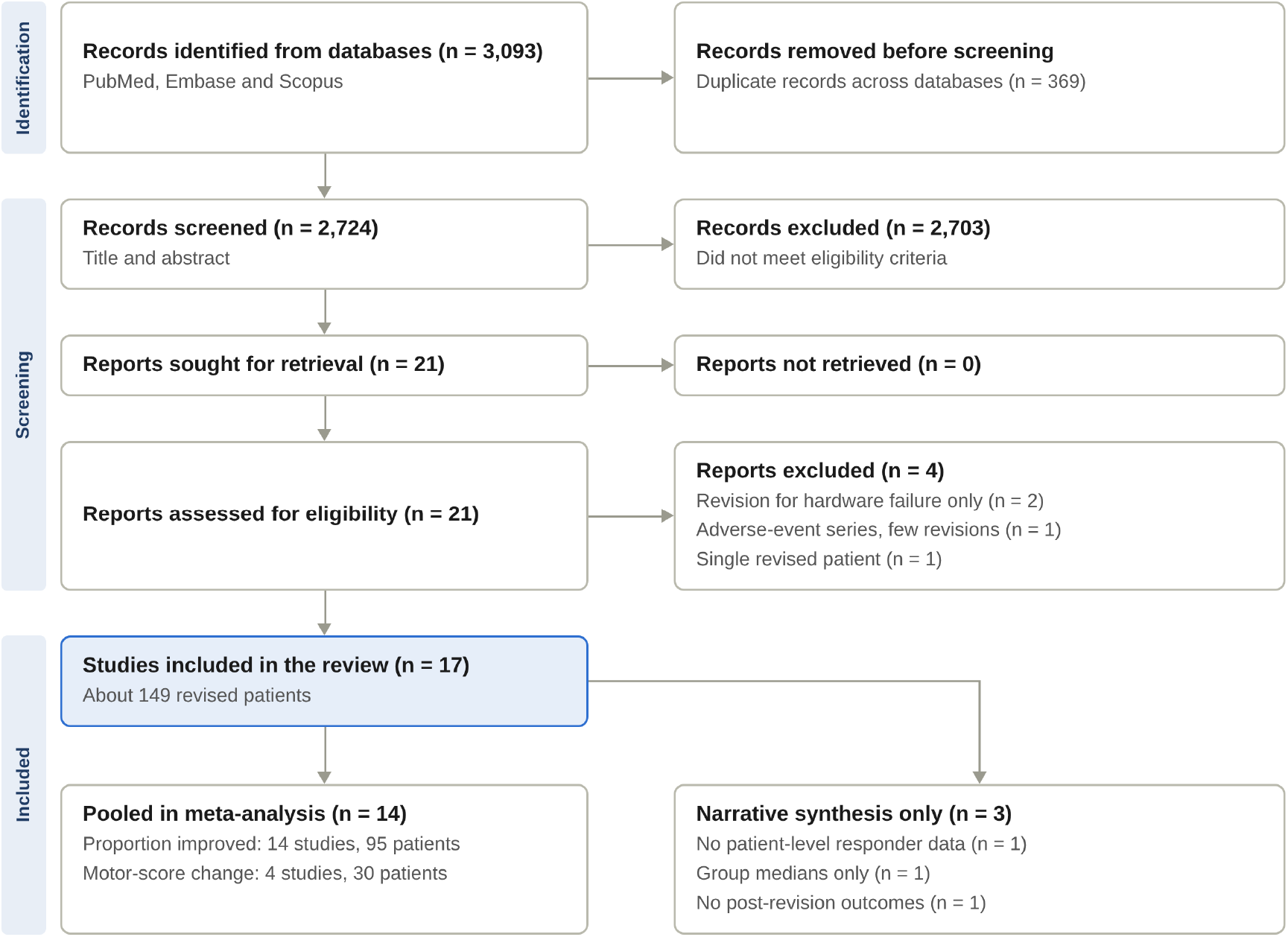
PRISMA 2020 flow diagram of study selection. Flow of records through identification, screening and inclusion, following the PRISMA 2020 template. Records excluded at title and abstract did not meet the eligibility criteria; the profile of the screened records is given in Supplementary Table S7, and the four full-text exclusions are listed in Supplementary Table S1.

The studies were published between 2005 and 2024 [8,9,33–47]. Seven came from the United States, two each from France and Germany, and one each from Japan, Sweden, Italy, Australia, China and a Canadian trial cohort. Eight were retrospective series, four were prospective or prospectively collected series, and five were small case series of two to four patients. Sample sizes ranged from 2 to 32 revised patients, approximately 149 in total (Table 1; some overlap between reports is possible). Eight studies concerned Parkinson disease, three essential tremor, three dystonia or dystonic tremor, and three mixed indications.

**Table 1.** Characteristics of the included studies.

| # | Study (year), country | Design | DBS indication and number revised | Revision type | Indication for revision |
|---|---|---|---|---|---|
| 1 | Okun 2005, USA | Retrospective case series, 2 centres | 41 referred "failures" (PD 31, ET 9, dystonia 1 at referral); 10 leads reimplanted | Lead reimplantation | Suboptimal outcome; 19/41 had misplaced leads, 10 reimplanted |
| 2 | Ellis 2008, USA | Retrospective case series, single centre | 22 patients, 25 leads: PD 11, ET 7, dystonia 4 (11 implanted in-house, 11 referred) | Lead repositioning or replacement | Suboptimal clinical benefit (infection and hardware cases excluded) |
| 3 | Anheim 2008, France | Prospective follow-up, single centre | 7 PD (12 of 14 electrodes misplaced); 5 bilateral, 2 unilateral | STN lead reimplantation | Poor response to STN DBS with misplacement on imaging |
| 4 | Richardson 2009, USA | Retrospective case series | 8 PD, 9 leads | STN lead repositioning | Inadequate improvement (4), adverse effects (1), both (3) |
| 5 | Yu 2009, USA | Case series | 3 ET | Added a second parallel thalamic lead (dual-channel IPG) | Loss of tremor control with intolerable adverse effects at high voltage |
| 6 | Blomstedt 2012, Sweden | Retrospective case series | 5 ET (4 M, 1 F), left side | Vim lead replaced or supplemented by cZi lead | Early (2) or late (3) failure of Vim DBS |
| 7 | Cif 2012, France | Open-label case series | 16 primary generalised dystonia (8 DYT1+) | Staged second pair of GPi electrodes | <50% improvement at 1 year, or later worsening, with optimal first leads |
| 8 | Miyagi 2013, Japan | Case series | 2 paediatric DYT1 dystonia | GPi lead replacement (case 1); added bilateral STN leads (case 2) | Early loss of benefit (tolerance) despite readjustment |
| 9 | Minafra 2014, Italy | Case series | 3 early-onset PD | Rescue GPi DBS 8 y after STN DBS | Worsening off periods, dystonia and freezing despite programming and medication changes |
| 10 | Cook 2015, Australia | Case series | 2 PD | Additional bilateral GPi DBS after STN DBS | Refractory disabling dyskinesia |
| 11 | Falowski 2016†, USA | Retrospective case series | 32 patients, 44 lead revisions from 497 leads (PD, ET, dystonia) | Repositioning, target change, added lead, replacement | Loss of efficacy (32 leads), adverse effects (7), malfunction (5) |
| 12 | Frizon 2019, USA | Retrospective case series | 25 patients/34 leads removed (PD); suboptimal-outcome subgroup 7 patients, 9 leads | Removal then reimplantation (STN; 2 STN→GPi, 1 STN→VIM) | Suboptimal outcome (infection and device failure analysed separately) |
| 13 | Nickl 2019, Germany | Consecutive prospectively collected series | 9 PD, 15 leads | STN lead replacement | DBS/levodopa response ratio <75% with lead >2 mm outside dorsolateral STN |
| 14 | Avecillas-Chasin 2022, Canada | Prospective trial participants, secondary analysis | 6 ET, 12 leads | Standard to directional lead replacement | Tremor progression with side effects limiting stimulation |
| 15 | Paschen 2023, Germany | Case series | 4 dystonic tremors with cervical dystonia | Rescue lead in the alternative target (GPi→Vim in 2; Vim→GPi in 2) | Insufficient response to first target |
| 16 | Zeng 2024, China | Retrospective case series | 6 PD (8 identified, 2 excluded) | GPi leads removed, STN leads implanted | Unsatisfactory efficacy of GPi DBS |
| 17 | Schnurman 2024, USA | Retrospective case series | 7 PD of 670 treated (1.0%) | Rescue lead: GPi (4), VIM (3) | Persistent dyskinesia (4) or residual tremor (3) after STN DBS |

PD, Parkinson disease; ET, essential tremor; STN, subthalamic nucleus; GPi, globus pallidus internus; Vim/VIM, ventral intermediate nucleus; cZi, caudal zona incerta; IPG, implantable pulse generator. † No post-revision clinical outcome scores reported.

Three revision strategies emerged (Table 2). Same-target revision (repositioning, replacement or reimplantation) was reported in nine studies, usually for a lead judged to be outside the intended target on imaging. Added or rescue leads were used in seven studies, in the same target (parallel or second pair of pallidal leads) or a different one (pallidal leads after subthalamic stimulation, thalamic leads for residual tremor). Target switching, for example from the pallidum to the subthalamic nucleus or from the thalamus to the caudal zona incerta, was described in four studies. Interval from the index operation to revision ranged from weeks to more than eight years, and reported lead displacement after repositioning ranged from about 1.6 to 8.4 mm (mean vector distance 2.2–6.7 mm in the studies that gave it).

**Table 2.** Revision strategy, timing and lead displacement.

| Study | Time from index surgery to revision | Lead displacement | Surgical approach | Follow-up after revision |
|---|---|---|---|---|
| Okun 2005 | Not reported | Not reported | Reimplantation | Not reported |
| Ellis 2008 | Mean 28.9 mo (2–104); in-house 9.7 mo, referred 48 mo | Mean vector distance: PD 5.5 mm, ET 6.1 mm, dystonia 6.7 mm | Old device removed, repeat microelectrode mapping | Not reported |
| Anheim 2008 | Median 16.9 mo (12–23) | Contact–target distance 5.4 → 2.0 mm | Misplaced electrode left in place; 5 parallel microelectrodes, local anaesthesia | 12 mo |
| Richardson 2009 | Mean 18 mo (6–44) | Vector 2.3–4.9 mm (mean 4.1) | Old lead as reference, no microelectrode recording, fluoroscopy; old lead kept in 6 of 9 | Not reported |
| Yu 2009 | Years of benefit before loss | New lead 2 mm anterior and medial to the original | Same burr hole, shared cap, dual-channel IPG | 36, 12 and 10 mo |
| Blomstedt 2012 | Early (2 patients) or years (3) | Vim and cZi leads 2.7–6.8 mm apart | New burr hole, local anaesthesia, macrostimulation | 12–24 mo |
| Cif 2012 | Mean 5.0 y | Second lead posterior in 28 GPi, anterior in 4 | Same burr hole | Mean 4.2 y after addition |
| Miyagi 2013 | Case 1: 1 wk, then at recurrence | Case 1: lateral replacement | Replacement; case 2 added STN leads | Case 1: years; case 2: died 2 d after reoperation |
| Minafra 2014 | 8 y after STN | New target | STN stimulation off after 3 mo | 24 mo |
| Cook 2015 | 1 y (case 1); 5.5 y (case 2) | New target | Separate burr holes lateral to STN entries, microelectrode recording | Not reported |
| Falowski 2016 | Mostly <1 y | STN leads moved mainly medial and/or anterior | Old lead kept in place during mapping and final placement | ≥6 mo |
| Frizon 2019 | Mean 32 d between removal and reimplantation (suboptimal subgroup) | 2.73 mm (SD 1.31); overall 2.16 mm | Staged removal then reimplantation, awake electrophysiology | Mean 36.6 mo (SD 26.3) |
| Nickl 2019 | Median 3 y (0.5–10) | Median 4.1 mm (1.6–8.4) | New entry point; old lead as marker; MER in all; 2 staged at 3 mo | Median 97 d (78–123) |
| Avecillas-Chasin 2022 | Mean 4.9 y (2.9–8.8) | 9 of 12 leads in same AP/ML position; 8 of 12 deeper | Same tract; directional lead | 6–16 wk |
| Paschen 2023 | 5–12 mo between targets | New target | Staged bilateral implantation | 3–8 mo after each step |
| Zeng 2024 | Mean 19.8 mo (10–32) | New target (GPi→STN) | Same burr holes | Mean 8.8 mo |
| Schnurman 2024 | Median 4.1 y (0.6–12.6) | New target | Separate burr hole; generator changed to dual-channel | Not reported |

AP, anteroposterior; ML, mediolateral; MER, microelectrode recording; mo, months; y, years; wk, weeks; d, days.

### Clinical outcomes

Table 3 lists the outcomes by study.

**Table 3.** Clinical outcomes after revision.

| Study | Primary outcome measure | Result |
|---|---|---|
| Okun 2005 | Clinician-judged outcome | Whole cohort: good 21/41 (51%), modest 6 (15%), none 14 (34%). Reimplanted leads: text reports marked improvement in 7 and partial in 3; the patient-level table shows 6 marked, 1 partial, 2 some, 1 none |
| Ellis 2008 | UPDRS-III, TRS, UDRS, CGIS, PDQ-39 | PD off-medication/on-DBS UPDRS-III −24.4% (n=6); ET TRS −60.4% (n=5); dystonia UDRS −12.8% (n=3). CGIS much or very much improved: PD 10/11, ET 7/7, dystonia 3/4 |
| Anheim 2008 | Off-medication/on-stimulation UPDRS-III | Improvement vs. basal state 26.7% → 59.4% at 1 y; median LEDD 1202 → 534 mg; Schwab & England 51% → 76%; 6/7 improved |
| Richardson 2009 | Chart-based functional improvement | All 8 improved; no rating-scale data |
| Yu 2009 | WHIGET tremor score | Pre-rescue → both leads: patient 1 12/14 → 12/14 (adverse effects severe → mild); patient 2 13/12 → 11/10; patient 3 5 → 2 |
| Blomstedt 2012 | ETRS hand tremor/function | Improvement 25% with Vim vs. 57% with cZi DBS; late failures did worse |
| Cif 2012 | BFMDRS motor score | Significant improvement after added electrodes: whole group p=0.005, DYT1− p=0.012, DYT1+ not significant |
| Miyagi 2013 | BFMDRS-M | Case 1: nadir 4.5, then recurrence to 63.5; no benefit from later replacements. Case 2: 78 → 15 at 3 mo, then 88 after added STN leads |
| Minafra 2014 | UPDRS-III, UPDRS-IV, LEDD (24 mo) | Off-medication UPDRS-III unchanged; UPDRS-IV 15→6, 13→10, 13→6; painful dystonia resolved in 2 |
| Cook 2015 | UPDRS-III, UPDRS-IV | Patient 1: dyskinesia suppressed, UPDRS-III 47 → 25 with GPi on. Patient 2: improved dyskinesia and levodopa tolerance |
| Falowski 2016 | None | Not reported |
| Frizon 2019 | MDS-UPDRS-III | Suboptimal subgroup 37.88 → 24.38 (p=0.025); after infection no difference (p=0.742) |
| Nickl 2019 | UPDRS-III off-medication/on-stimulation; DBS/levodopa response ratio | 38.2 ± 6.6 → 15.5 ± 7.9 (p<0.001); response ratio 64% → 190% |
| Avecillas-Chasin 2022 | Tremor improvement (FTM-TRS) | Median 48.7% with standard DBS → 68.4% with directional DBS (p=0.017) |
| Paschen 2023 | FTM-TRS and UDRS | Extremity tremor (GPi first): UDRS −45%, tremor unchanged; after Vim rescue FTM-TRS −51%. Axial tremor (Vim first): FTM-TRS −24%; after GPi rescue −63%, UDRS −49% |
| Zeng 2024 | Off-medication MDS-UPDRS-III | −37.5% vs. baseline (p=0.002), −29.2% vs. GPi DBS (p=0.030); 4/6 responders |
| Schnurman 2024 | Dyskinesia/tremor grade, UPDRS-III | All 4 GPi rescues improved dyskinesia; all 3 VIM rescues improved tremor |

CGIS, Clinician Global Impression Scale; WHIGET, Washington Heights–Inwood Genetic Study of Essential Tremor; ETRS, essential tremor rating scale; BFMDRS-M, Burke–Fahn–Marsden Dystonia Rating Scale motor score; FTM-TRS, Fahn–Tolosa–Marin Tremor Rating Scale; UDRS, Unified Dystonia Rating Scale; LEDD, levodopa equivalent daily dose.

#### Parkinson disease, same-target revision

After subthalamic lead repositioning, the off-medication/on-stimulation motor score improved by 59% from baseline at 1 year in the prospective French series, compared with 27% before reimplantation, and levodopa-equivalent dose fell from 1,202 to 534 mg [34]. In the German series, the motor score fell from 38.2 to 15.5 and the stimulation-to-levodopa response ratio rose from 64% to 190%, and stimulation-induced adverse effects resolved in all nine patients [9]. The Cleveland series found a fall from 37.9 to 24.4 points after revision for suboptimal outcome (p=0.025) but no change after reimplantation for infection [43]. Richardson and colleagues reported functional improvement in all eight patients, without rating-scale data [35], and Ellis and colleagues a 24.4% improvement in motor score in six patients with Parkinson disease [33].

#### Parkinson disease, rescue leads and target switching

Adding a pallidal lead for dyskinesia after subthalamic stimulation improved dyskinesia in all four patients in the largest series [47], both patients in an Australian report [41], and two of three in an Italian report [40]. Thalamic rescue leads improved residual tremor in all three patients [47]. Switching from pallidal to subthalamic stimulation after unsatisfactory pallidal outcome gave a 37.5% improvement in off-medication motor score versus baseline in six patients, with four responders [46].

#### Essential tremor

In five evaluable patients, repeat thalamic lead placement reduced tremor score by 60.4% [33]. Moving to the caudal zona incerta improved hand tremor and function by 57% compared with 25% on the failed ventral intermediate lead, but late failures did less well than early ones [37]. A second parallel lead recaptured tremor control or tolerability in three patients [36], and replacement with directional leads raised median tremor improvement from 48.7% to 68.4% [44].

#### Dystonia and dystonic tremor

Adding a second pair of pallidal electrodes improved the Burke–Fahn–Marsden motor score significantly in 16 patients with primary generalised dystonia (p=0.005), but not in the DYT1-positive subgroup [38]. Two children with DYT1 dystonia derived no durable benefit from lead replacement or added subthalamic leads [39]. In four patients with dystonic tremor and cervical dystonia, a second target rescued the outcome in each (tremor score −51% and −63%), with the right target depending on tremor distribution [45]. Ellis and colleagues saw a 12.8% improvement in dystonia scores [33].

### Pooled analyses

Fourteen studies (95 patients) contributed to the pooled proportion (Figure 2; Supplementary Tables S2 and S3). Three studies could not be pooled: one lacked patient-level responder data [38], one reported group medians only [44], and one gave no post-revision outcomes [42]. Meaningful improvement occurred in 81% (95% CI 68–91%; I² 50%, Q p=0.016). Estimates were 82% (64–95%; I² 64%) for same-target revision (6 studies, 63 patients) and 79% (59–93%; I² 41%) for added, rescue or switched leads (8 studies, 32 patients). Counting any improvement, the pooled proportion was 92% (84–97%; I² 30%). Leave-one-out estimates ranged from 79% to 84%, excluding studies with fewer than five patients gave 82% (69–93%), and counting the inconsistent study from its text gave 82% (70–91%).

**Figure 2.**
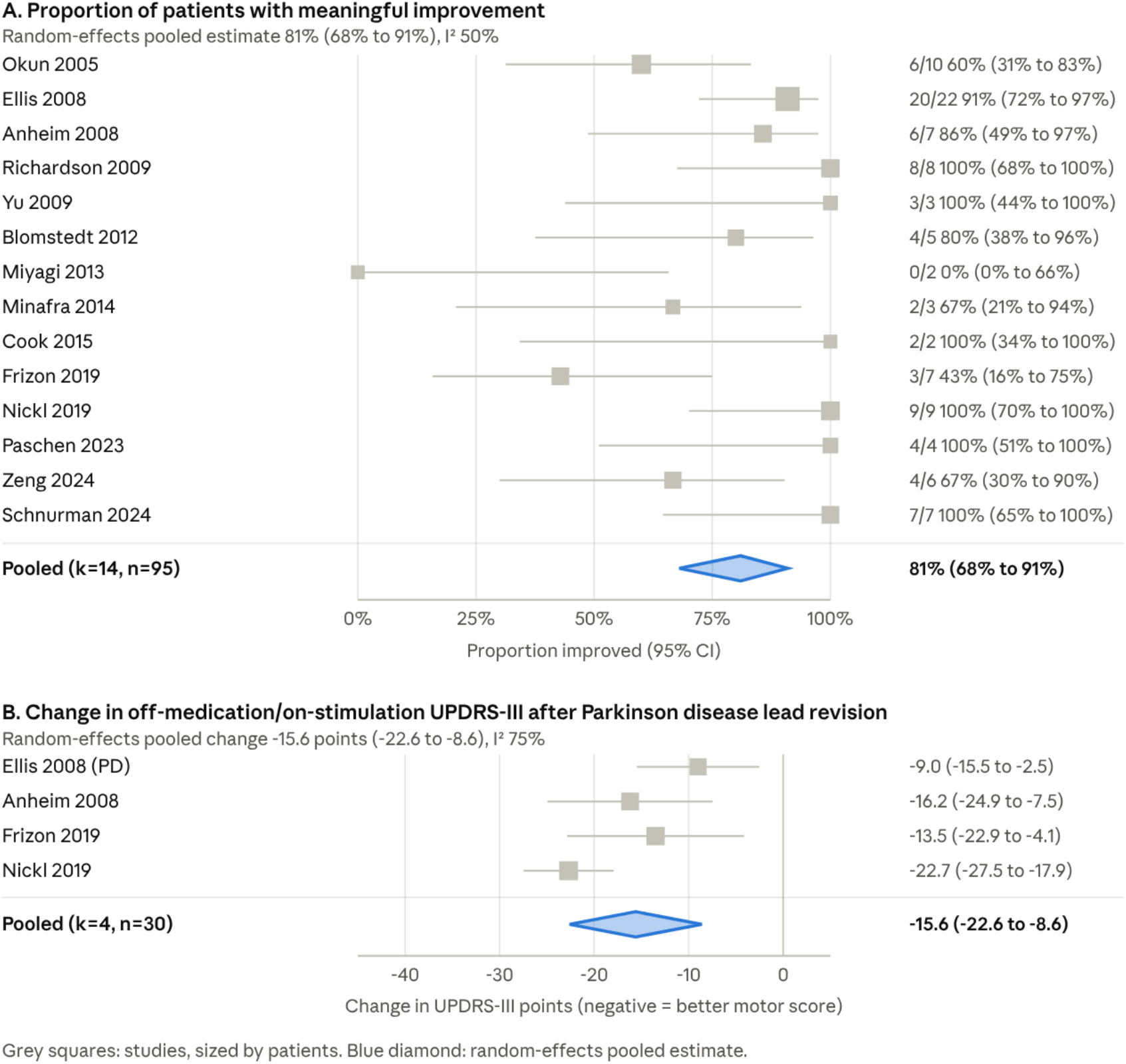
Forest plots of the pooled analyses. (A) Proportion of patients with meaningful improvement in each of 14 studies, with Wilson 95% confidence intervals and the random-effects pooled estimate (diamond). (B) Change in off-medication/on-stimulation UPDRS-III in four studies of lead revision in Parkinson disease; standard deviations of change were calculated from raw data, derived from reported p values or imputed assuming a pre–post correlation of 0.5. Squares are sized by the number of patients.

Four studies of subthalamic or Parkinson lead revision (30 patients) reported comparable off-medication/on-stimulation motor scores [9,33,34,43]. The pooled change was −15.6 points (95% CI −22.6 to −8.6; I² 75%, τ² 37.2; Hartung–Knapp interval −25.3 to −6.0). Varying the assumed correlation between 0.3 and 0.7 changed the estimate by 0.3 points.

### Complications

Seven studies reported at least one surgical or hardware complication, eight reported none, and two did not report complications separately for revised patients (Table 4). Reported events included infection requiring hardware removal (two patients in revision series [33,34]), lead or extension fractures, skin erosion or poor wound healing, and a thalamic hemorrhage of less than 5 mL in a patient who had a pulmonary embolism, who later died of medical causes [43]. A second death occurred in a child with DYT1 dystonia and status dystonicus, 2 days after added subthalamic leads and during emergency surgery [39]. Stimulation-related adverse effects (dysarthria, paranesthesia, diplopia, depression, ataxia) resolved after revision in most reports that tracked them [9,34,35,44].

**Table 4.** Complications and adverse events related to revision.

| Study | Surgical and hardware events | Stimulation-related adverse effects |
|---|---|---|
| Okun 2005 | Not reported for the revised subgroup | Not reported |
| Ellis 2008 | Infection requiring hardware removal (1); lead fracture, connector wire replaced (1); friable skin with exposed hardware, converted to thalamotomy (1) | Not reported |
| Anheim 2008 | Transient infection, stimulator removed, reimplanted 2 mo later, no sequelae (1) | Threshold for first adverse effect 2.6 → 4.4 V |
| Richardson 2009 | None | Speech impairment, facial contractions, depression and diplopia resolved |
| Yu 2009 | None described | Paraesthesia and dysarthria reduced to mild or none |
| Blomstedt 2012 | None described | Headache and sensory effects with Vim stimulation (1) |
| Cif 2012 | 7 hardware events over follow-up (3 before, 4 after the second pair): 2 infections, 2 extension fractures, 1 electrode fracture, 1 IPG dysfunction, 1 migration; no haemorrhage | Not reported |
| Miyagi 2013 | 2 lead fractures (case 1); case 2: death 2 d after reoperation (oedema, heart failure, status dystonicus, rhabdomyolysis) | Not reported |
| Minafra 2014 | None reported | Not reported |
| Cook 2015 | None | Not reported |
| Falowski 2016 | 1 skin erosion; no cranial wound infections | Adverse effects were an indication in 7 leads |
| Frizon 2019 | Poor wound healing after reimplantation (1); left thalamic haemorrhage (<5 mL) after pulmonary embolism, death at another facility (1) | Not reported |
| Nickl 2019 | None | Dysarthria, hypomania/impulsivity and ataxia resolved |
| Avecillas-Chasin 2022 | Not reported | Pre-revision dysarthria (5), gait problems (2), paraesthesia (1) |
| Paschen 2023 | Prolonged wound healing without sequelae (1) | Dysarthria or gait ataxia with Vim stimulation (1) |
| Zeng 2024 | No severe or persistent adverse events | Not reported |
| Schnurman 2024 | No operative complications | All patients tolerated dual stimulation |

### Factors associated with success or failure

The included studies point to five recurring factors (Table 5). First, **lead position**: benefit was greatest when the lead was moved into the central dorsolateral STN, and the distance from the active contact to the theoretical target correlated inversely with improvement (p=0.02) [34,35]. Revision was effective even for small displacements, with a median of 4.1 mm in the German series [9]. Second, **preserved levodopa response** and a stimulation-to-levodopa response ratio below 75% identified patients likely to benefit [9,34]. Third, **timing and course**: in essential tremor, early failures did better than late deterioration [37], and late loss of benefit in dystonia was often resistant to revision [38,39]. Fourth, **phenotype**: persistent dyskinesia responded to pallidal and residual tremor to thalamic rescue leads [47], and in dystonic tremor the better target depended on whether tremor was extremity-predominant or axial [45]. Fifth, **age and genotype**: DYT1 status and early childhood onset were associated with poor or short-lived benefit [38,39].

**Table 5.** Factors associated with revision success or failure.

| Study | Factors reported |
|---|---|
| Okun 2005 | Misplaced lead on imaging; wrong diagnosis and dementia gave poor results; 51% salvaged by revision, programming and medication |
| Ellis 2008 | Asymmetric benefit and lead migration as clues; referred leads had longer latency (48 vs. 9.7 mo) |
| Anheim 2008 | Smaller contact–target distance correlated with greater improvement (Spearman p=0.02); preserved levodopa response |
| Richardson 2009 | Central dorsolateral STN effective; peripheral leads ineffective; lateral leads gave corticobulbar effects, medial leads speech effects |
| Yu 2009 | Target between two adjacent leads; new lead alone no better than original |
| Blomstedt 2012 | Early failures responded better than late; greater baseline tremor linked to more residual tremor; fibrous sheet around removed leads |
| Cif 2012 | DYT1− improved, DYT1+ did not; DYT1+ patients younger at first surgery (12.9 vs. 21.1 y) |
| Miyagi 2013 | Correctly placed GPi leads; early childhood age raised as possible tolerance factor |
| Minafra 2014 | Preserved levodopa response and correct STN lead position; axial and dyskinetic features |
| Cook 2015 | Young-onset disease; dyskinesia refractory to medication and STN adjustment |
| Falowski 2016 | Most revisions within 1 year; STN leads usually moved medial/anterior |
| Frizon 2019 | Benefit after suboptimal-outcome revision but not after infection-related reimplantation; limited intraoperative physiology at index surgery in 2 patients |
| Nickl 2019 | DBS/levodopa response ratio <75% with image-confirmed lead >2 mm outside dorsolateral STN; preserved levodopa response |
| Avecillas-Chasin 2022 | Directional steering; more ventral (posterior subthalamic area) position; pallidofugal and cerebellothalamic pathways |
| Paschen 2023 | Tremor distribution: extremity-predominant responded to Vim, axial-predominant to GPi |
| Zeng 2024 | Low initial improvement vs. levodopa response; persistent rigidity and bradykinesia |
| Schnurman 2024 | Young age at onset (median 33.5 y); persistent dyskinesia responded to GPi, residual tremor to VIM |

### Risk of bias and certainty of evidence

On the JBI checklist, 16 of 17 studies scored 70% or more (Supplementary Table S4), but the checklist does not capture the main threats. No study had a comparison group, most assessors were unblinded (blinded video rating was used in three studies and partly in a fourth), and all 17 studies carried moderate (6 studies) or high (11 studies) concern in the broader assessment (Supplementary Table S5). Certainty of evidence was very low for clinical improvement, motor-score change, complications and predictors (Supplementary Table S6).

## Discussion

### Principal findings

This is, to our knowledge, the first systematic review to focus on surgical revision for suboptimal DBS outcomes. In 17 case series covering approximately 149 patients, most revised patients improved: 81% had meaningful benefit, and patients with Parkinson disease who underwent subthalamic lead revision had a mean reduction of 15.6 points in the off-medication/on-stimulation motor score. Same-target revision and added or switched leads performed similarly. The evidence is nonetheless of very low certainty, and the pooled figures should be read as a description of published revised patients, not as an estimate of how often revision works.

### Interpretation of the pooled estimates

Several features push the pooled results upward. There is no comparison group, so regression to the mean and continuing programming are indistinguishable from a surgical effect. Patients who improve are more likely to be written up, and the proportions are close to 100% in many small series. Outcome criteria differ between studies and were mostly applied by the treating team. The 11-percentage-point gap between “any improvement” (92%) and “meaningful improvement” (81%) shows that some benefit is modest. Heterogeneity was moderate for proportions (I² 50%) and high for motor score change (I² 75%), and the two pooled analyses rest on 95 and 30 patients. We therefore regard the analysis as exploratory.

### Mechanisms of benefit and failure

The consistent message of the Parkinson series is that small spatial errors matter. Leads that lay outside the dorsolateral STN gave partial benefit and adverse effects at low voltage, and moving them by about 2–5 mm produced large gains [9,34,35,43]. Lead location also relates to outcome in essential tremor [13]. In Anheim’s series the distance between the active contact and the theoretical target fell from 5.4 to 2.0 mm, and improvement correlated with proximity [34]. That a median displacement of 4.1 mm was enough in the German series [9] implies that revision is worth considering even when a lead looks acceptable on routine imaging, provided stimulation response fails to match the levodopa response. Directional leads and short pulse widths may handle part of this problem without surgery [16,17], which is why exhaustive reprogramming comes first. In referral cohorts, inadequate programming explained more than a third of apparent failures [8], and only 9 of 48 referred patients remained suboptimal after reprogramming [9]; revision candidates are therefore a minority.

A second group of patients had correctly placed leads and a clear but incomplete response. Here revision meant adding a second lead to address the residual symptom, with a pallidal lead for dyskinesia and a thalamic lead for tremor [40,41,47]. This extends the rescue-lead strategy introduced in small reports [18,19]. Because the added lead leaves the original in place, it keeps the benefit already obtained and avoids the risk of losing it, which is the main worry of lead replacement.

Revision did not help when the problem lay in the disease or in the brain’s response to stimulation. Tolerance and disease progression may explain why late-onset loss of benefit in essential tremor [37] and early loss in children with DYT1 dystonia [39] responded poorly, whatever the lead position. The benefit of revision was lowest in these patients, and in children an additional lead was followed by a fatal course in one [39]. These patients are probably better served by other options, such as medication, stimulation holidays or a different target chosen after case review, than by repeat surgery on the same circuit.

### A proposed pathway for revision

The studies suggest, but do not prove, a stepwise approach. (1) Confirm the diagnosis and exclude hardware failure by impedance checks and radiographs [10,11]. (2) Verify lead position on fused imaging against the intended target. (3) Exhaust reprogramming, including directional and short-pulse options. (4) Benchmark stimulation against levodopa response in Parkinson disease [9,48]. (5) Choose the strategy: reposition when the lead is demonstrably outside the target and the response is partial, add a rescue lead when the lead is well placed and a specific symptom persists, and consider a target switch when the initial target has stopped working. (6) Decide in a multidisciplinary team, and discuss with the patient that tolerance, progression and non-responders limit what surgery can offer. This pathway is a hypothesis for testing, not a guideline.

### Surgical technique

Surgeons used either the existing lead as an internal reference and removed it late or left it in place [35,44], or removed it first and implanted after a delay [9,43]. Some used microelectrode recording and awake testing, others did not [35,37]. Comparative data are absent, so we cannot say whether staged or single-step revision gives better placement. Staged revision avoids tracking into the old channel but leaves the patient without stimulation for weeks.

### Safety

Most series reported few complications, but event counts are small and often unreported. Infection, lead fracture and wound problems were the commonest events [33,34,38,43]. Two deaths occurred in approximately 149 patients, one of cardiopulmonary origin after a small hemorrhage [43] and one in a child in status dystonicus [39]. The expected gain from revision should be weighed against these risks and against the possibility of losing the partial benefit of an existing lead.

### Strengths and limitations

Strengths include a focus on suboptimal outcome rather than hardware failure, full-text data extraction with study-level subgroup extraction, a pre-defined appraisal framework, transparent definitions of improvement and sensitivity analyses. The main limitations reflect the evidence base: the included studies are small, uncontrolled and mostly retrospective, with probable publication bias. The pooled proportion rests on study-specific, mostly unblinded definitions, and standard deviations for the continuous outcome were partly imputed. Two groups of studies may share patients, so effective sample size may be smaller. Studies mixing indications forced subgroup extraction, and heterogeneity across disease, target and technique limits generalization. Our search was restricted to published English-language studies in three bibliographic databases and the reference lists of included studies, so grey literature, trial registries, non-English reports (84 records at screening) and unindexed reports may have been missed, and study authors were not contacted for missing data. We could not assess publication bias statistically.

### Implications for research

The field needs a prospective, multicenter registry of revisions with pre-specified indications, uniform outcome measures (for instance the stimulation-to-levodopa response ratio in Parkinson disease and a blinded tremor or dystonia scale), imaging-verified lead positions and reporting of failures. Trials comparing repositioning with reprogramming using new technologies, and rescue leads with target switching, are feasible in high-volume centers. Reporting of negative and null cases would help correct the bias in the present literature.

## Conclusions

For selected patients with suboptimal DBS outcomes despite optimized programming, published experience shows clear improvement in most patients after surgical revision, with infrequent but sometimes serious complications. Revision is most promising when imaging shows a lead outside the target and the response is still partial, or when a well-placed lead leaves a specific symptom uncontrolled. It is least promising for late tolerance and in young children with DYT1 dystonia. Because the evidence is very low in certainty, revision should be decided case by case within a multidisciplinary team until prospective data are available.

## Declarations

### Author contributions

MAS is the guarantor of the review also, conceptualization, search, screening, data extraction, risk-of-bias appraisal, screening, data extraction, original draft, writing, review and editing.

SB: formal analysis, writing, search, screening, data extraction, original draft, writing, review and editing.

### Ethics

Not applicable (systematic review of published data).

### Data availability

The extraction sheets and analysis code are available from the corresponding author on reasonable request.

### Registration

PROSPERO CRD420261557397. The full search strategy was uploaded with the record.

### Online supplementary material

- **Supplementary Appendix 1**. Search strategies. **Supplementary Table S1.** Studies excluded at full text. **S2.** Datasets for the pooled analyses. **S3.** Pooled and sensitivity results. **S4.** JBI checklist. **S5.** Additional bias domains. **S6.** Certainty of evidence. **S7.** Profile of the records screened.

## Data Availability

All data produced in the present study are available upon reasonable request to the authors

## Supplementary material

### Supplementary Appendix 1. Search strategies

Databases: PubMed, Embase and Scopus, from inception to October 2026, with no date or language limits in the search (English-language studies were selected at screening). We only putted the PubMed Search string and the main search query with most data retrieved. The full strategy for other Databases was uploaded with the PROSPERO record (https://www.crd.york.ac.uk/PROSPEROFILES/e0f26240ae1ac4a5a6d6c06254fb6c16.pdf).

### PubMed

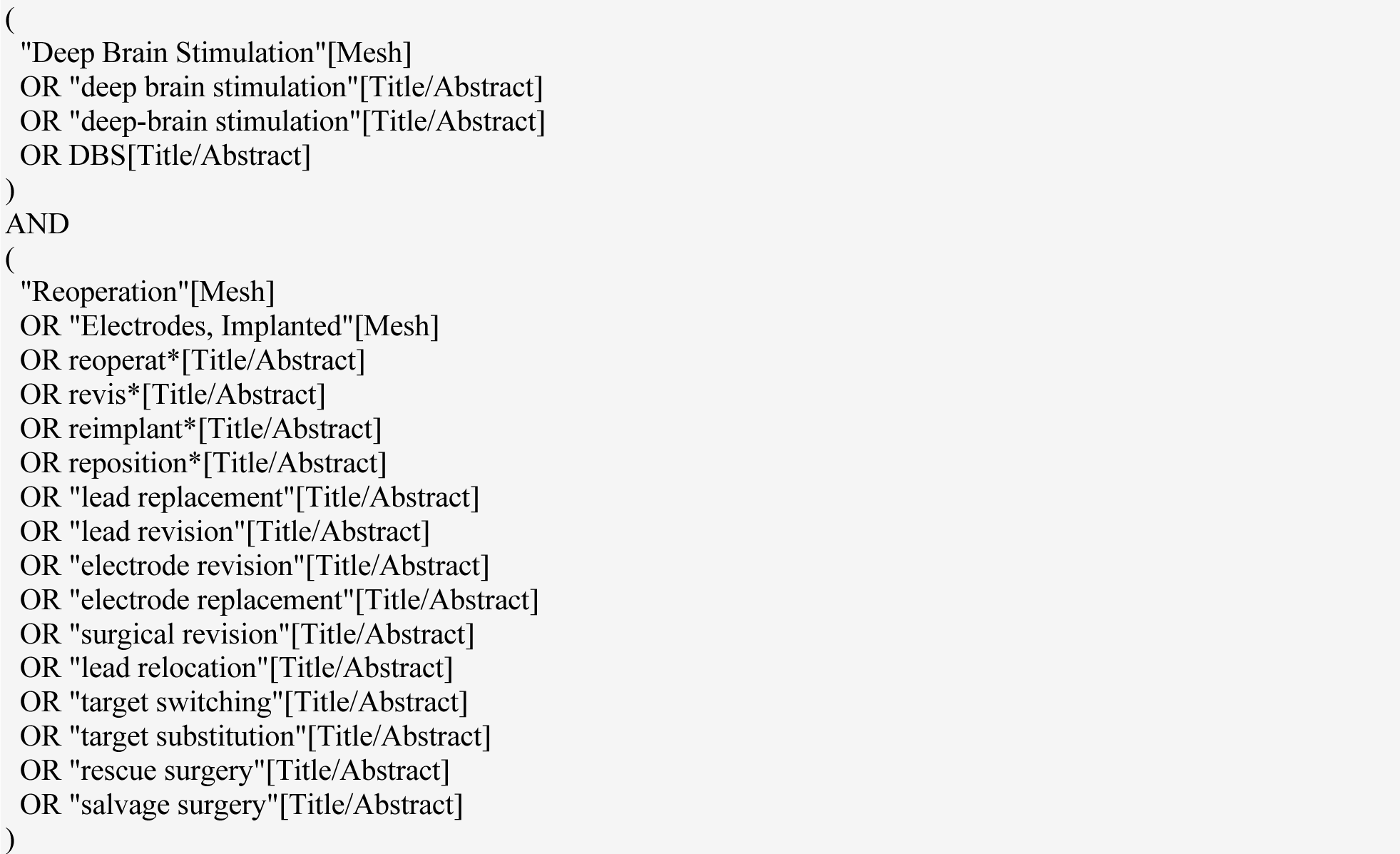

**Supplementary Table S1.** Studies excluded at the full-text stage.

| Study | Reason for exclusion |
|---|---|
| Guridi 2012 [10] | Revision for hardware failure (lead fracture or disruption with impedance rise), not for suboptimal outcome |
| Allert 2011 [11] | Hardware dysfunction study (short circuits, disconnections, loose contacts); revision in 4 of 52 dysfunctional electrodes, mainly managed by reprogramming |
| Nonaka 2020 [31] | General adverse-event series with few revision cases |
| Anthofer 2017 [32] | Tractography distance study with a single revised patient |

**Supplementary Table S2a.** Dataset for the pooled proportion.

| Study | Group | n | Meaningful improvement | Any improvement | Definition used |
|---|---|---|---|---|---|
| Okun 2005 | A | 10 | 6 | 9 | Marked improvement (meaningful); marked, partial or some (any), from the patient-level table |
| Ellis 2008 | A | 22 | 20 | 22 | CGIS 1–2 (meaningful); CGIS 1–3 (any) |
| Anheim 2008 | A | 7 | 6 | 6 | All but one patient improved |
| Richardson 2009 | A | 8 | 8 | 8 | Authors: significant functional improvement in each |
| Yu 2009 | B | 3 | 3 | 3 | Authors: tremor control recaptured with fewer adverse effects |
| Blomstedt 2012 | B | 5 | 4 | 5 | ≥50% lower hand score with cZi on vs. off (calculated); any improvement |
| Miyagi 2013 | B | 2 | 0 | 0 | No sustained benefit after replacement or added leads |
| Minafra 2014 | B | 3 | 2 | 3 | UPDRS-IV and painful dystonia improved in 2; marginal in 1 |
| Cook 2015 | B | 2 | 2 | 2 | Dyskinesia suppressed |
| Frizon 2019 | A | 7 | 3 | 7 | ≥25% reduction in motor score (calculated); any reduction |
| Nickl 2019 | A | 9 | 9 | 9 | All improved; adverse effects resolved |
| Paschen 2023 | B | 4 | 4 | 4 | Authors: significant benefit from the second target |
| Zeng 2024 | B | 6 | 4 | 4 | ≥24% improvement vs. GPi DBS (any improvement not given per patient) |
| Schnurman 2024 | B | 7 | 7 | 7 | Dyskinesia or tremor improved |

Group A, replacement or repositioning within the same target. Group B, added, rescue or switched lead or target (Miyagi, mixed, placed in B). Not pooled: Cif 2012 (no patient-level responder data), Avecillas-Chasin 2022 (group medians only), Falowski 2016 (no post-revision outcomes).

**Supplementary Table S2b.** Dataset for the pooled UPDRS-III change.

| Study | n | Before | After | Mean change | SD of change (source) |
|---|---|---|---|---|---|
| Ellis 2008 (6 PD, 1 GPi) | 6 | 36.8 | 27.8 | −9.0 | 8.1 (raw patient data) |
| Anheim 2008 | 7 | 40.1 (median) | 22.2 (median) | −16.2 | 11.8 (imputed, r=0.5; means estimated from medians and ranges) |
| Frizon 2019 (suboptimal subgroup) | 8 | 37.88 ± 8.78 | 24.38 ± 13.09 | −13.5 | 13.5 (derived from p=0.025; n inferred) |
| Nickl 2019 | 9 | 38.2 ± 6.6 | 15.5 ± 7.9 | −22.7 | 7.3 (imputed, r=0.5) |

UPDRS-III and MDS-UPDRS-III are treated as the same scale in points.

**Supplementary Table S3a.**
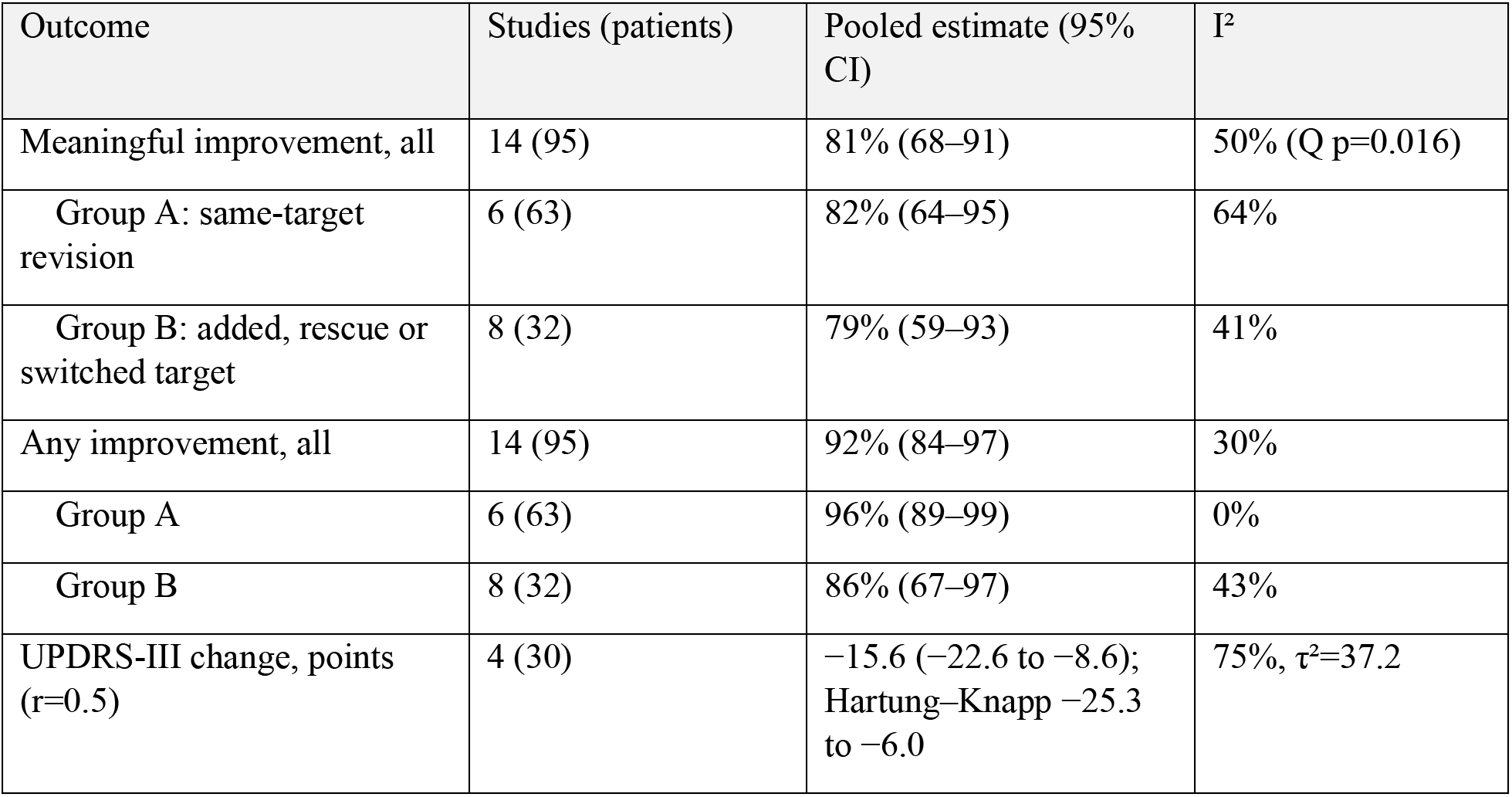
Pooled results.

| Outcome | Studies (patients) | Pooled estimate (95% CI) | I<sup>2</sup> |
|---|---|---|---|
| Meaningful improvement, all | 14 (95) | 81% (68–91) | 50% (Q p=0.016) |
| Group A: same-target revision | 6 (63) | 82% (64–95) | 64% |
| Group B: added, rescue or switched target | 8 (32) | 79% (59–93) | 41% |
| Any improvement, all | 14 (95) | 92% (84–97) | 30% |
| Group A | 6 (63) | 96% (89–99) | 0% |
| Group B | 8 (32) | 86% (67–97) | 43% |
| UPDRS-III change, points (r=0.5) | 4 (30) | −15.6 (−22.6 to −8.6); Hartung–Knapp −25.3 to −6.0 | 75%, τ<sup>2</sup>=37.2 |

**Supplementary Table S3b.**
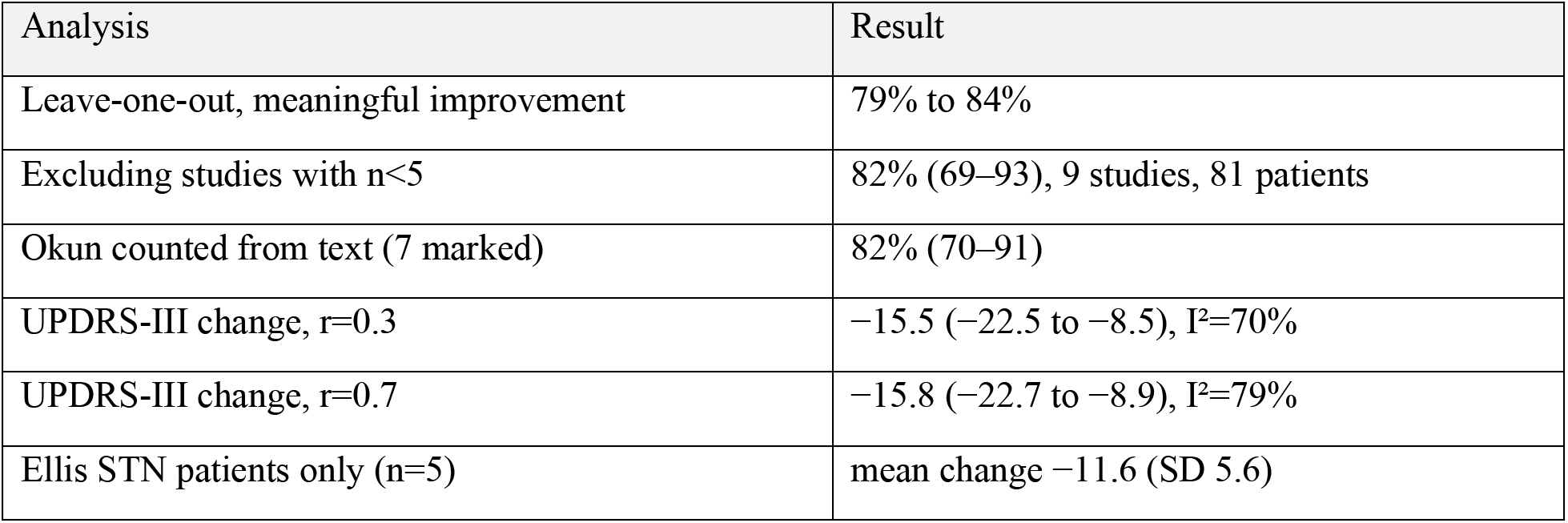
Sensitivity analyses.

| Analysis | Result |
|---|---|
| Leave-one-out, meaningful improvement | 79% to 84% |
| Excluding studies with n<5 | 82% (69–93), 9 studies, 81 patients |
| Okun counted from text (7 marked) | 82% (70–91) |
| UPDRS-III change, r=0.3 | −15.5 (−22.5 to −8.5), I<sup>2</sup>=70% |
| UPDRS-III change, r=0.7 | −15.8 (−22.7 to −8.9), I<sup>2</sup>=79% |
| Ellis STN patients only (n=5) | mean change −11.6 (SD 5.6) |

**Supplementary Table S4.** JBI checklist for case series.

| Study | Q1 | Q2 | Q3 | Q4 | Q5 | Q6 | Q7 | Q8 | Q9 | Q10 | Yes / applicable | Rating |
|---|---|---|---|---|---|---|---|---|---|---|---|---|
| Okun 2005 | Y | Y | Y | Y | Y | Y | Y | U | Y | NA | 8/9 (89%) | Low |
| Ellis 2008 | Y | Y | Y | U | Y | Y | Y | U | Y | NA | 7/9 (78%) | Low |
| Anheim 2008 | Y | Y | Y | Y | Y | Y | Y | Y | Y | Y | 10/10 (100%) | Low |
| Richardson 2009 | Y | Y | Y | U | Y | Y | Y | U | Y | NA | 7/9 (78%) | Low |
| Yu 2009 | Y | Y | Y | U | Y | Y | Y | Y | Y | NA | 8/9 (89%) | Low |
| Blomstedt 2012 | Y | Y | Y | U | Y | Y | Y | U | Y | NA | 7/9 (78%) | Low |
| Cif 2012 | Y | Y | Y | Y | Y | Y | Y | Y | Y | Y | 10/10 (100%) | Low |
| Miyagi 2013 | Y | Y | Y | Y | Y | Y | Y | Y | Y | NA | 9/9 (100%) | Low |
| Minafra 2014 | Y | Y | Y | U | Y | Y | Y | Y | Y | NA | 8/9 (89%) | Low |
| Cook 2015 | Y | Y | Y | U | Y | Y | Y | Y | Y | NA | 8/9 (89%) | Low |
| Falowski 2016 | Y | Y | Y | Y | Y | U | U | N | Y | NA | 6/9 (67%) | Moderate |
| Frizon 2019 | Y | Y | Y | Y | Y | Y | Y | U | Y | Y | 9/10 (90%) | Low |
| Nickl 2019 | Y | Y | Y | Y | Y | Y | Y | Y | Y | Y | 10/10 (100%) | Low |
| Avecillas-Chasin 2022 | Y | Y | Y | U | Y | Y | Y | Y | Y | Y | 9/10 (90%) | Low |
| Paschen 2023 | Y | Y | Y | U | Y | Y | Y | Y | Y | Y | 9/10 (90%) | Low |
| Zeng 2024 | Y | Y | Y | U | Y | Y | Y | Y | Y | Y | 9/10 (90%) | Low |
| Schnurman 2024 | Y | Y | Y | Y | Y | Y | Y | U | Y | NA | 8/9 (89%) | Low |

Q1 clear inclusion criteria; Q2 condition measured in a standard, reliable way; Q3 valid identification methods; Q4 consecutive inclusion; Q5 complete inclusion; Q6 demographics reported; Q7 clinical information reported; Q8 outcomes or follow-up clearly reported; Q9 setting reported; Q10 appropriate statistics. Y, yes; N, no; U, unclear; NA, not applicable (descriptive study). Cut-offs: ≥70% low, 50–69% moderate, <50% high. Ratings are preliminary.

**Supplementary Table S5.** Additional bias domains and overall concern.

| Study | Outcome scale | Assessor blinded | Main concerns | Overall concern |
|---|---|---|---|---|
| Okun 2005 | Clinician global judgement | No | Referral selection; mixed interventions; subjective outcome | High |
| Ellis 2008 | UPDRS-III, TRS, UDRS, CGIS | No | Some scores missing; overlap with Okun possible | High |
| Anheim 2008 | UPDRS-III | Not stated | Small n; no control | Moderate |
| Richardson 2009 | None (chart review) | No | No rating scales; small n | High |
| Yu 2009 | WHIGET | Independent neurologists, not blinded to lead status | 3 patients | High |
| Blomstedt 2012 | ETRS | No | Non-systematic follow-up; evaluated by the treating team | High |
| Cif 2012 | BFMDRS-M | Not stated | No control; added leads confound disease progression | Moderate |
| Miyagi 2013 | BFMDRS-M | No | 2 patients | High |
| Minafra 2014 | UPDRS-III, IV | No | 3 patients; STN stopped at 3 mo | High |
| Cook 2015 | UPDRS, video | No | 2 patients | High |
| Falowski 2016 | None | No | No post-revision outcomes | High |
| Frizon 2019 | MDS-UPDRS-III | No | Scores missing for several leads; mixed indications | High |
| Nickl 2019 | UPDRS-III | Not stated | Small n; no control | Moderate |
| Avecillas-Chasin 2022 | FTM-TRS | Yes (blinded video rating) | 6 patients; patients also reported elsewhere | Moderate |
| Paschen 2023 | FTM-TRS, UDRS | Yes (blinded video rating) | 4 patients; fixed order of targets | Moderate |
| Zeng 2024 | MDS-UPDRS-III | Yes (blinded video) | 6 patients; 8.8 mo follow-up; 2 of 8 excluded | Moderate |
| Schnurman 2024 | UPDRS, dyskinesia grade | Partly (2 videos) | Retrospective; subjective dyskinesia; missing scores | High |

**Supplementary Table S6.**
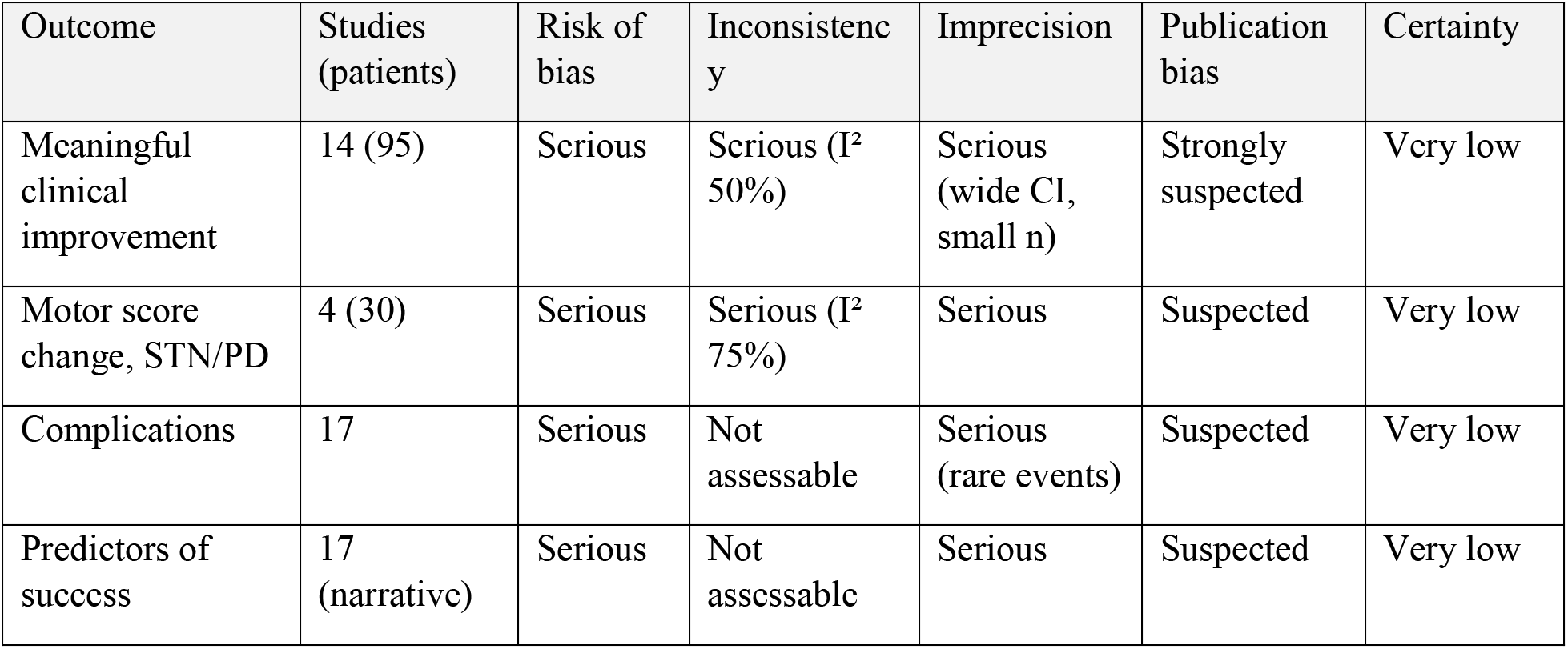
Certainty of evidence (GRADE-style)

| Outcome | Studies (patients) | Risk of bias | Inconsistency | Imprecision | Publication bias | Certainty |
|---|---|---|---|---|---|---|
| Meaningful clinical improvement | 14 (95) | Serious | Serious (I² 50%) | Serious (wide CI, small n) | Strongly suspected | Very low |
| Motor score change, STN/PD | 4 (30) | Serious | Serious (I² 75%) | Serious | Suspected | Very low |
| Complications | 17 | Serious | Not assessable | Serious (rare events) | Suspected | Very low |
| Predictors of success | 17 (narrative) | Serious | Not assessable | Serious | Suspected | Very low |

**Supplementary Table S7.** Profile of the 2,724 unique records screened.

| Category | Records, n | % of 2,724 |
|---|---|---|
| Indexed as human studies | 2,339 | 85.9 |
| Animal-only (indexed as animals, not humans) | 226 | 8.3 |
| Human status unknown (not yet MeSH-indexed) | 131 | 4.8 |
| Randomized trials | 51 | 1.9 |
| Other clinical trials | 116 | 4.3 |
| Case reports | 383 | 14.1 |
| Systematic reviews | 46 | 1.7 |
| Meta-analyses | 22 | 0.8 |
| Other (narrative or overview) reviews | 316 | 11.6 |
| No abstract | 161 | 5.9 |
| Non-English | 84 | 3.1 |

Categories come from database indexing (MeSH and publication type) and are not mutually exclusive. The search returned 3,093 records, of which 369 duplicates across databases were removed before screening.

